# Disruption of the CCL5/RANTES-CCR5 Pathway Restores Immune Homeostasis and Reduces Plasma Viral Load in Critical COVID-19

**DOI:** 10.1101/2020.05.02.20084673

**Authors:** Bruce K. Patterson, Harish Seethamraju, Kush Dhody, Michael J. Corley, Kazem Kazempour, Jay Lalezari, Alina P.S. Pang, Christopher Sugai, Edgar B. Francisco, Amruta Pise, Hallison Rodrigues, Mathew Ryou, Helen L. Wu, Gabriela M. Webb, Byung S. Park, Scott Kelly, Nader Pourhassan, Alena Lelic, Lama Kdouh, Monica Herrera, Eric Hall, Enver Aklin, Lishomwa C. Ndhlovu, Jonah B. Sacha

## Abstract

Severe acute respiratory syndrome coronavirus 2 (SARS-CoV-2), the causative agent of coronavirus disease 2019 (COVID-19), is now pandemic with nearly three million cases reported to date^1^. Although the majority of COVID-19 patients experience only mild or moderate symptoms, a subset will progress to severe disease with pneumonia and acute respiratory distress syndrome (ARDS) requiring mechanical ventilation^2^. Emerging results indicate a dysregulated immune response characterized by runaway inflammation, including cytokine release syndrome (CRS), as the major driver of pathology in severe COVID-19^3,4^. With no treatments currently approved for COVID-19, therapeutics to prevent or treat the excessive inflammation in severe disease caused by SARS-CoV-2 infection are urgently needed. Here, in 10 terminally-ill, critical COVID-19 patients we report profound elevation of plasma IL-6 and CCL5 (RANTES), decreased CD8+ T cell levels, and SARS-CoV-2 plasma viremia. Following compassionate care treatment with the CCR5 blocking antibody leronlimab, we observed complete CCR5 receptor occupancy on macrophage and T cells, rapid reduction of plasma IL-6, restoration of the CD4/CD8 ratio, and a significant decrease in SARS-CoV-2 plasma viremia. Consistent with reduction of plasma IL-6, single-cell RNA-sequencing revealed declines in transcriptomic myeloid cell clusters expressing IL-6 and interferon-related genes. These results demonstrate a novel approach to resolving unchecked inflammation, restoring immunologic deficiencies, and reducing SARS-CoV-2 plasma viral load via disruption of the CCL5-CCR5 axis, and support randomized clinical trials to assess clinical efficacy of leronlimab-mediated inhibition of CCR5 for COVID-19.

## MAIN TEXT

Since the initial cases of COVID-19 were reported from Wuhan, China in December 2019^2^, SARS-CoV-2 has emerged as a global pandemic with an ever-increasing number of severe cases requiring invasive external ventilation that threatens to overwhelm health care systems^1^. While it remains unclear why COVID-19 patients experience a spectrum of clinical outcomes ranging from asymptomatic to severe disease, the salient features of COVID-19 pathogenesis and mortality are rampant inflammation and CRS leading to ARDS^4,5^. Indeed, excessive immune cell infiltration into the lung, cytokine storm, and ARDS have previously been described as defining features of severe disease in humans infected with the closely related betacoronaviruses SARS-CoV and MERS-CoV^6,7^. Because SARS-CoV-infected airway epithelial cells and macrophages express high levels of CCL5^8,9^, a chemotactic molecule able to amplify inflammatory responses towards immunopathology, we hypothesized that disrupting the CCL5-CCR5 axis via leronlimab-mediated CCR5 blockade would prevent pulmonary trafficking of pro-inflammatory leukocytes and reverse cytokine storm in COVID-19.

Leronlimab, formerly PRO 140, is a CCR5-specific human IgG4 monoclonal antibody in development for HIV therapy as a once-weekly, at-home subcutaneous injection. In five completed and four ongoing HIV clinical trials where over 800 individuals have received leronlimab, no drug related deaths, serious injection site reactions, or drug-drug interactions were reported^10–13^. Self-administration of leronlimab by patients facilitates simple, once-weekly dosing. In contrast to the small molecule CCR5 inhibitors that prevent HIV Env binding to CCR5 via allosteric modulation, leronlimab binds to the CCR5 extracellular loop 2 domain and N-terminus, thereby directly blocking the binding of HIV Env to the CCR5 co-receptor via a competitive mechanism. Leronlimab does not downregulate CCR5 surface expression or deplete CCR5-expressing cells, but does prevent CCL5-induced calcium mobilization in CCR5+ cells with an IC_50_ of 45 μg/ml^14^. This ability to specifically prevent CCL5-induced activation and chemotaxis of inflammatory CCR5+ macrophages and T cells suggests that leronlimab might be effective in resolving pathologies involving the CCL5-CCR5 pathway.

Ten critical COVID-19 patients at the Montefiore Medical Center received leronlimab via FDA-approved emergency investigational new drug (EIND) requests for individual patient use (Table 1). These confirmed SARS-CoV-2 positive patients had significant pre-existing co-morbidities and were receiving intensive care treatment including mechanical ventilation or supplemental oxygen for ARDS. Consistent with previous reports of severe COVID-19 disease^2^, these patients showed evidence of lymphopenia with liver and kidney damage (Supplementary Fig. 1)^15^. Four of the patients died during the fourteen-day study period due to a combination of disease complications and severe constraints on medical equipment culminating in medical triage. Although this EIND study lacks a placebo control group for comparison, a recent study of other critically ill COVID-19 patients in the New York City area indicates mortality rates as high as 88%^16^.

**Table 1:**
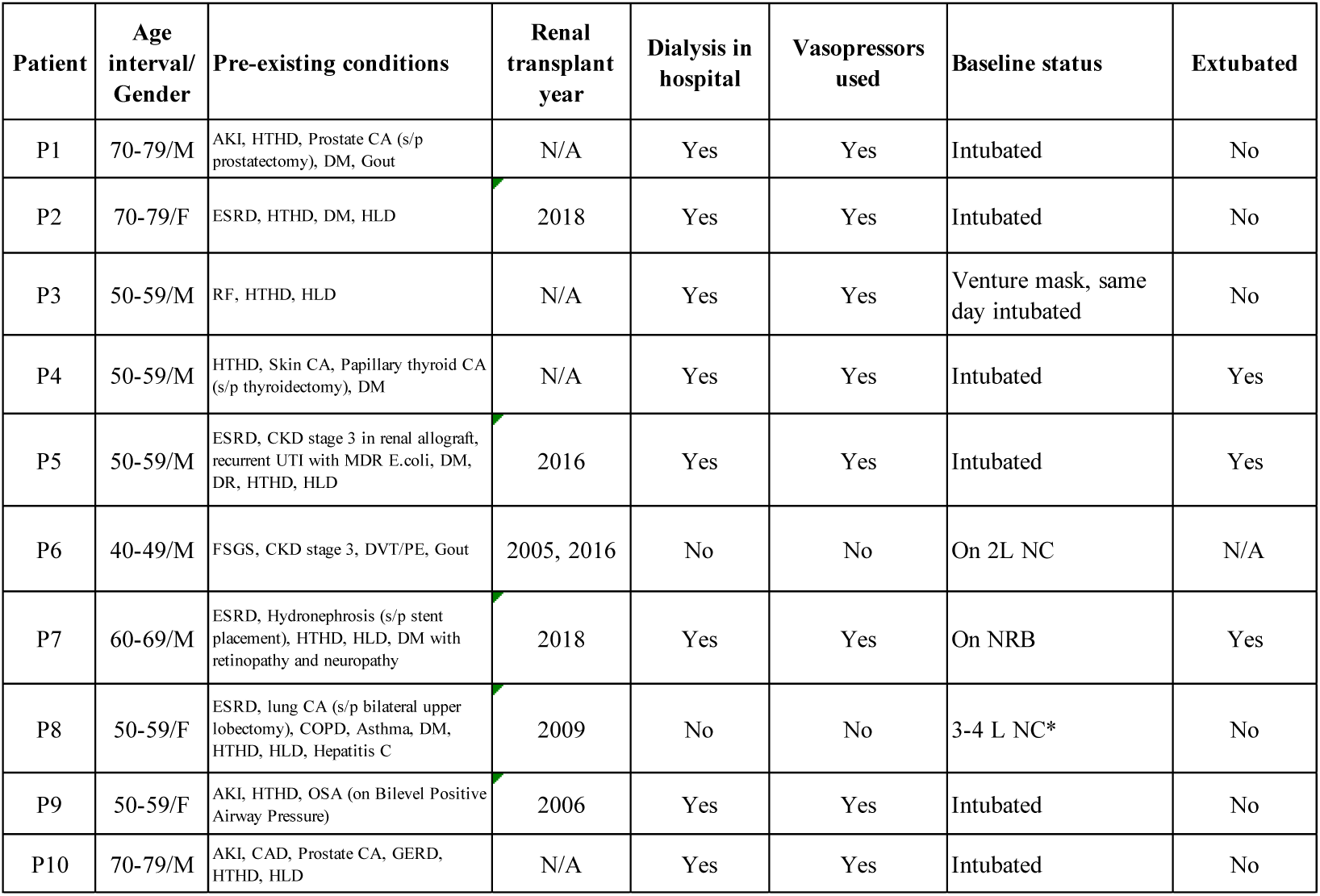
Critical COVID-19 Patient Summaries. N/A = not applicable, s/p = status post-, AKI = acute kidney injury, HTHD = hypertensive heart disease, DM = diabetes mellitus, HLD = hyperlipidemia, ESRD = end-stage renal disease, HD = hemodialysis, CA = cancer, COPD = chronic obstructive pulmonary disease, LUL = left upper lobe, RUL = right upper lobe, MDR = multi-drug resistant, CKD = chronic kidney disease, UTI = urinary tract infection, FSGS = Focal segmental glomerulosclerosis, DVT = deep vein thrombosis, PE = pulmonary embolism, OSA = obstructive sleep apnea, CAD = coronary artery disease, GERD = gastroesophageal reflux disease, RF = renal failure, DR = diabetic retinopathy, NC = nasal canula, NRB = non-rebreather mask, *Patient declined intubation due to poor baseline pulmonary status.

| Patient | Age interval/<br>Gender | Pre-existing conditions | Renal transplant year | Dialysis in hospital | Vasopressors used | Baseline status | Extubated |
| --- | --- | --- | --- | --- | --- | --- | --- |
| P1 | 70-79/M | AKI, HTHD, Prostate CA (s/p prostatectomy), DM, Gout | N/A | Yes | Yes | Intubated | No |
| P2 | 70-79/F | ESRD, HTHD, DM, HLD | 2018 | Yes | Yes | Intubated | No |
| P3 | 50-59/M | RF, HTHD, HLD | N/A | Yes | Yes | Venture mask, same day intubated | No |
| P4 | 50-59/M | HTHD, Skin CA, Papillary thyroid CA (s/p thyroidectomy), DM | N/A | Yes | Yes | Intubated | Yes |
| P5 | 50-59/M | ESRD, CKD stage 3 in renal allograft, recurrent UTI with MDR E.coli, DM, DR, HTHD, HLD | 2016 | Yes | Yes | Intubated | Yes |
| P6 | 40-49/M | FSGS, CKD stage 3, DVT/PE, Gout | 2005, 2016 | No | No | On 2L NC | N/A |
| P7 | 60-69/M | ESRD, Hydronephrosis (s/p stent placement), HTHD, HLD, DM with retinopathy and neuropathy | 2018 | Yes | Yes | On NRB | Yes |
| P8 | 50-59/F | ESRD, lung CA (s/p bilateral upper lobectomy), COPD, Asthma, DM, HTHD, HLD, Hepatitis C | 2009 | No | No | 3-4 L NC* | No |
| P9 | 50-59/F | AKI, HTHD, OSA (on Bilevel Positive Airway Pressure) | 2006 | Yes | Yes | Intubated | No |
| P10 | 70-79/M | AKI, CAD, Prostate CA, GERD, HTHD, HLD | N/A | Yes | Yes | Intubated | No |

Hyper immune activation and cytokine storm are present in cases of severe COVID-19^4^. Indeed, at leronlimab treatment baseline, signatures of CRS were present in the plasma of all ten patients in the form of significantly elevated levels of the inflammatory cytokines IL-1β, IL-6, and IL-8 (Fig. 1a-c) compared to healthy controls. In comparison to patients with mild or moderate COVID-19, only IL-6 was present at significantly higher levels in critically-ill patients. Of note, plasma CCL5 levels in the ten critically ill patients were markedly elevated over those in both healthy controls and mild or moderate COVID-19 patients (Fig. 1d). High levels of CCL5 can cause acute renal failure and liver toxicity^17,18^, both common findings in COVID-19 infection. Indeed, the critically ill patients presented with varying degrees of kidney and liver injury, although many had also previously received kidney transplants^15^ (Table 1 and Supplementary Fig. 1).

**Figure 1.**
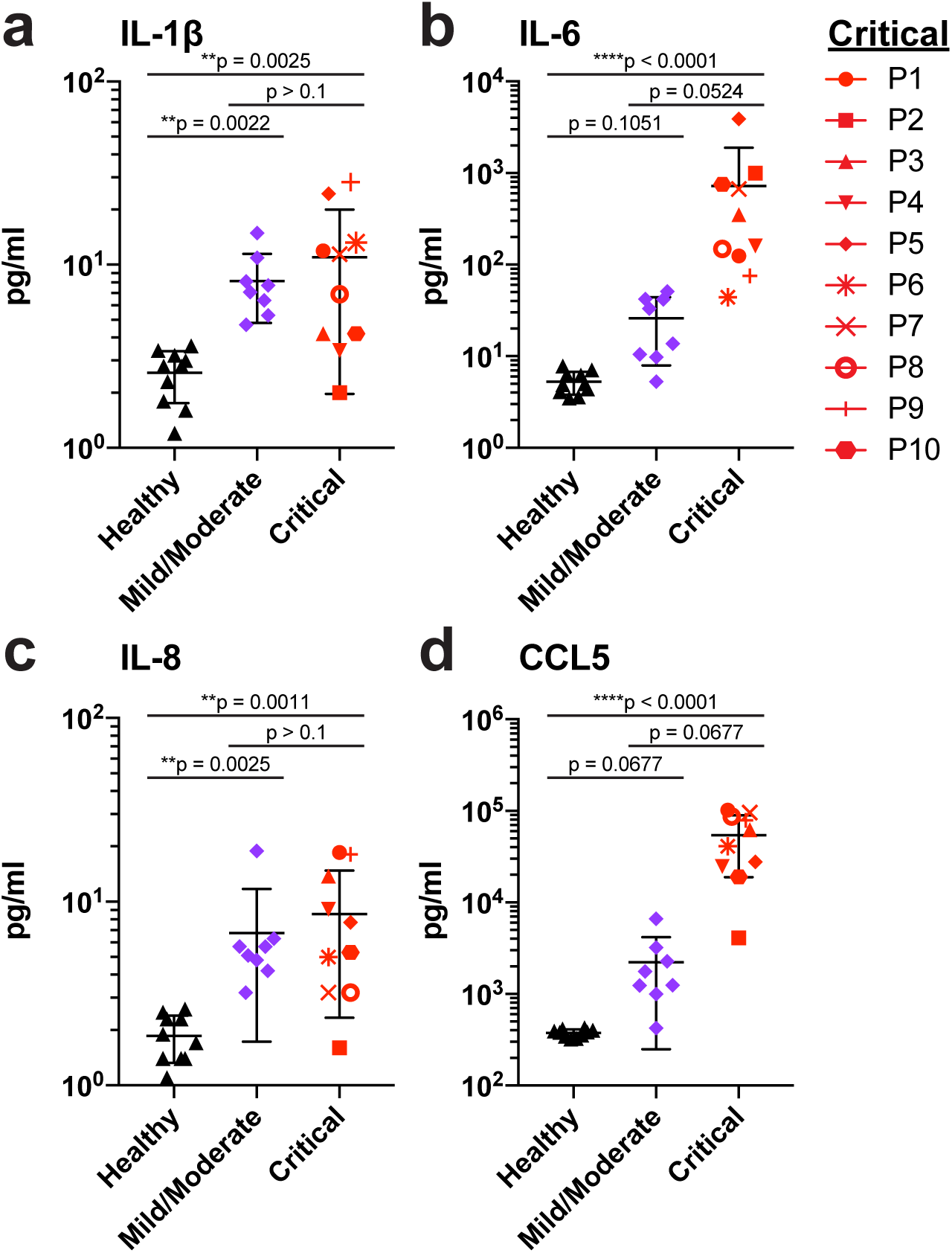
Elevated cytokine and chemokine levels in critically ill COVID-19 patients. **a-d**, Plasma levels of IL-1β (**a**), IL-6 (**b**), IL-8 (**c**), and CCL5 (**d**) in patients with mild/moderate (n=8, purple symbols) and critical (n=10, red symbols) COVID-19 disease, compared to healthy controls (n=10, black symbols). Graphs display p-values calculated by Dunn’s Kruskal-Wallis test: *p ≤ 0.05, ** p ≤ 0.01, ***p ≤ 0.001, ****p ≤ 0.0001.

At study day zero, all ten critically ill patients received a subcutaneous 700mg injection of leronlimab following baseline blood collection. Because defining features of severe COVID-19 disease include plasma IL-6 and T cell lymphopenia^2,19^, and we observed >100-fold increased CCL5 levels compared to normal controls (Fig. 1d), we longitudinally monitored these parameters for two weeks after leronlimab treatment. A reduction of plasma IL-6 was observed as early as three days following leronlimab and returned to healthy control levels by day 14 (Fig. 2a). In contrast, more variable levels were observed with IL-1β, IL-8, and CCL5 after leronlimab treatment (Supplementary Fig. 2). Following leronlimab administration, a marked restoration of CD8+ T cells (Fig. 2b) and a normalization of the CD4+ and CD8+ T cell ratio in blood was observed (Fig. 2c). These immunological changes occurred concomitant with full leronlimab CCR5 receptor occupancy on the surface of CCR5+ T cells and macrophages (Fig. 2d, 2e). Low levels of SARS-CoV-2 have been detected, but not yet quantified in the plasma of COVID-19 patients^19^. We used high sensitivity, digital droplet PCR to quantify plasma SARS-CoV-2 viremia at baseline. SARS-CoV-2 was found in the plasma of all ten critically ill patients, underscoring the severity of COVID-19 (Fig. 2f). Following leronlimab administration SARS-CoV-2 plasma viremia decreased in all patients at day seven, suggesting more effective anti-viral immunity following leronlimab-mediated CCR5 blockade.

**Figure 2.**
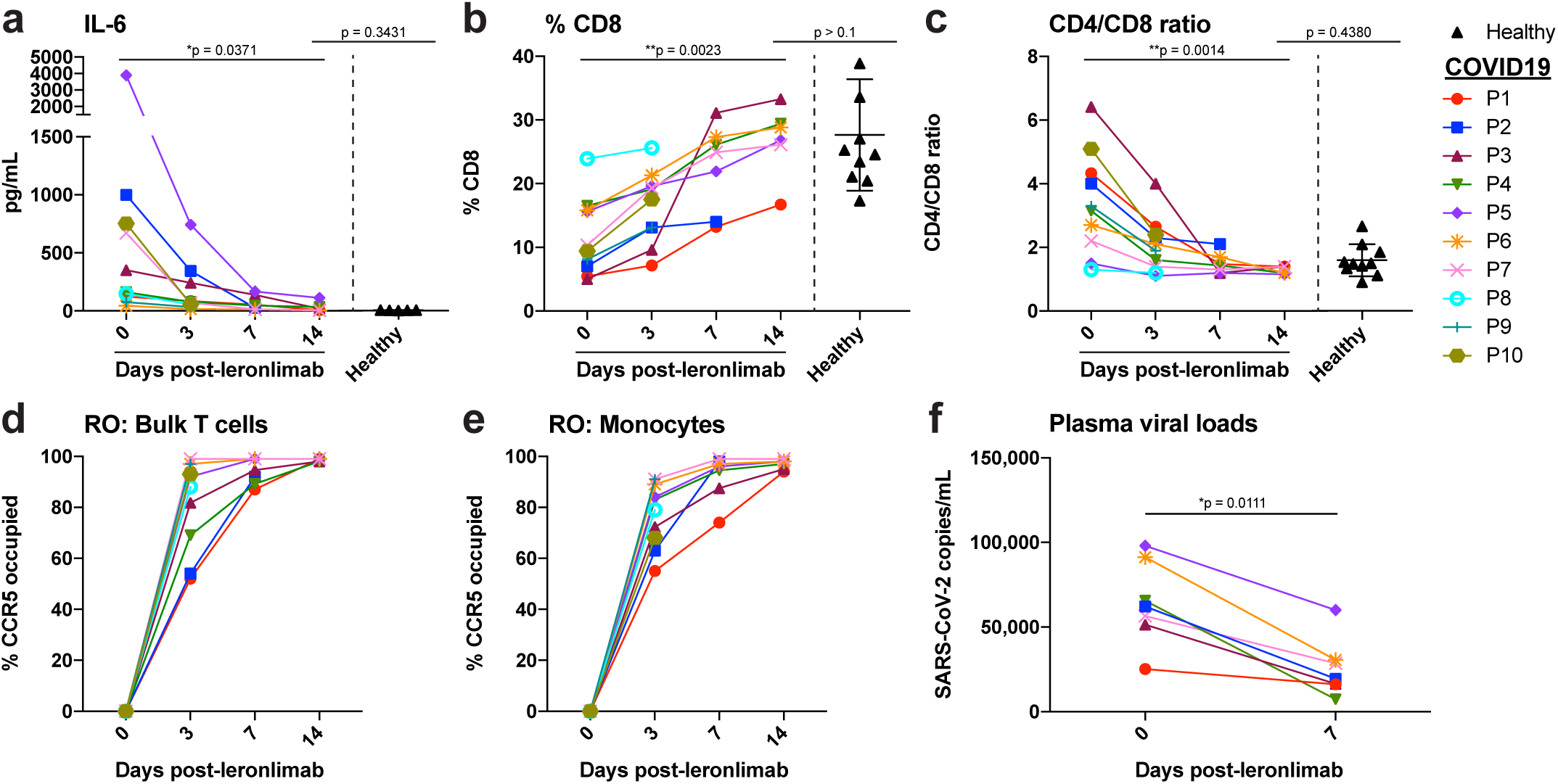
Reversal of immune dysfunction and CCR5 receptor occupancy in critically ill COVID-19 patients after leronlimab administration. **a-c**, Plasma levels of IL-6 (**a**), and peripheral blood CD8+ T cell percentages of CD3+ cells (**b**) and CD4/CD8 T cell ratio (**c**) at days 0 (n=10), 3 (n=10), 7 (n=7), and 14 (n=6) post-leronlimab administration. Healthy controls (n=10) shown in black triangles. Graphs display p-values calculated by Dunn’s Kruskal-Wallis test: not significant p > 0.05, *p ≤ 0.05, ** p ≤ 0.01, ***p ≤ 0.001, ****p ≤ 0.0001. **d-e**, CCR5 receptor occupancy on peripheral blood bulk T cells (**d**), and monocytes (**e**). **f**, SARS-CoV-2 plasma viral load at days 0 and 7 post-leronlimab (n=7). Graph displays p-value calculated by Mann-Whitney test: *p ≤ 0.05, ** p ≤ 0.01, ***p ≤ 0.001, ****p ≤ 0.0001.

Finally, to establish an unbiased gene repertoire for these COVID-19 patients, we performed 10X Genomics 5’ single cell RNA-sequencing of peripheral blood mononuclear cells to evaluate transcriptional changes between an uninfected healthy donor and two of the severe COVID-19 patients (P2 and P4) for which sufficient baseline, pre-leronlimab treatment COVID-19 samples were available for this analysis. We identified 2,890 differentially expressed transcripts between the two groups and found that the two severe COVID-19 patients had a greater abundance of myeloid cells upregulating inflammatory, interferon (IFN)-, and chemokine-related genes compared to a healthy control (FDR < 0.05) (Supplementary Table 2). Notable genes overexpressed in COVID-19 samples included chemokines (CXCL8, CCL4, CCL3), inflammatory and immune activation genes (IL-1b, CD69), and the IFN-related genes (IFI27, IFITM3) (Supplementary Fig. 3). We also observed a downregulation of the effector molecule granzyme A and the immunoregulatory gene KLRB1 compared to the healthy control.

To identify markers that would inform effective leronlimab treatment we conducted differential expression analysis for the same two severe COVID-19 participants (P2 and P4) for which baseline and day seven post leronlimab samples were available. Our longitudinal COVID-19 single cell dataset profiled an estimated 4,105 cells at baseline and 4,888 cells at the 7-day post leronlimab timepoint. We identified 2,037 differentially expressed transcripts (FDR < 0.05) (Supplementary Table 2). In line with the decrease of IL-6 protein levels observed in plasma, IL-6 transcripts were downregulated between day 0 and day 7 in monocytes, (Supplementary Fig. 4), consistent with reports of monocyte/macrophages repolarization following CCR5 blockade^20^. We observed that myeloid cells expressing chemokine and IFN-related genes such as CCL3, CCL4, CCL5, ADAR, APOBEC3A, IFI44L, ISG15, MX1 were downregulated at day 7 post leronlimab compared to baseline (Fig. 3 and Supplementary Table 3). Within the T cell population, we observed increased expression of granzyme A, suggesting improved antiviral function. These transcriptomic findings further underscore the potential impact of leronlimab-mediated CCR5 blockade on the inflammatory state in COVID-19.

**Figure 3.**
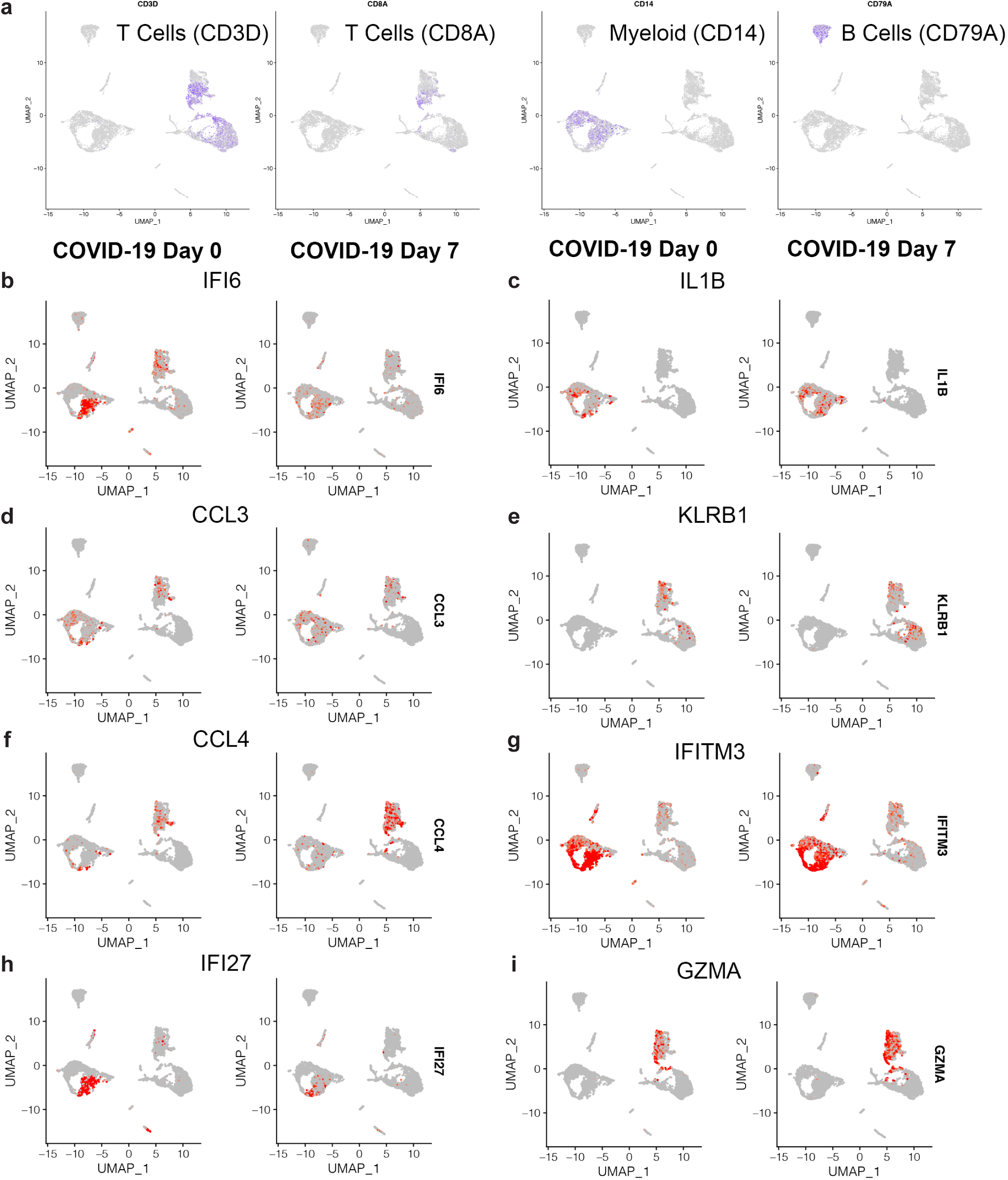
Longitudinal single-cell transcriptomics of COVID-19 following leronlimab. UMAP feature plots of single-cell transcriptome profiles of CD3 (T cells) versus CD8 (CD8+ T cells) versus CD14 (monocyte/myeloid) versus CD79a (B cells) (**a**) IFI6 (**b**), IL-1β (**c**), CCL3 (**d**), KLRB1 (**e**), CCL4 (**f**), IFITM3 (**g**), IFI27 (**h**), Granzyme A (**i**), before and 7 days post leronlimab treatment for severe COVID-19 patients P2 and P4.

Here, we report on the involvement of the CCL5-CCR5 pathway in COVID-19 and present data from ten critically ill patients with severe COVID-19 demonstrating reduction of inflammation, restoration of T cell lymphocytopenia, and reduced SARS-CoV-2 plasma viremia following leronlimab-mediated CCR5 blockade. Recent studies have found that a significant number of COVID-19 patients experience increased risks of strokes, blood clots and other thromboembolic events^21^. Platelet activation, which leads to the initiation of the coagulation cascade, can be triggered by chemokines including CCL5^22^, suggesting that leronlimab treatment may be beneficial beyond its immunomodulatory effects on inflammation and hemostasis in COVID-19 patients.

Given medical triage resulting in patient death, we cannot comment on the impact of leronlimab on clinical outcome in these patients. While anecdotal evidence of clinical improvement in COVID-19 patients following leronlimab treatment have been reported^23^, randomized controlled trials are required to determine efficacy of leronlimab for COVID-19. Indeed, randomized, double blind, placebo controlled clinical trials are underway to assess the efficacy of leronlimab treatments in patients with mild to moderate (NCT04343651)^24^ and severe to critical (NCT04347239)^25^ COVID-19. In summary, we show here for the first time, involvement of the CCL5-CCR5 axis in the pathology of SARS-CoV-2, and present evidence that inhibition of CCL5 activity via CCR5 blockade represents a novel therapeutic strategy for COVID-19 with both immunologic and virologic implications.

## METHODS

### Assessment of plasma cytokine and chemokine levels

Fresh plasma was used for cytokine quantification using a customized 13-plex bead-based flow cytometric assay (LegendPlex, Biolegend, Inc) on a CytoFlex flow cytometer. For each patient sample 25 μL of plasma was used in each well of a 96-well plate. Raw data was analyzed using LegendPlex software (Biolegend, Inc San Diego CA). Samples were run in duplicate. In addition, split sample confirmation testing was performed by ELISA (MDBiosciences, Minneapolis, MN). A 48-plex cytokine/chemokine/growth factor panel and RANTES-CCL5 (Millipore Sigma) assay were performed following manufacture’s protocol on a Luminex MAGPIX instrument. Confirmation testing was also performed in duplicate. Samples falling outside the linear range of the appropriate standard curves were diluted and repeated incorporating the dilution factor into the final average. Cytokine, chemokines and growth factors included: sCD40L, EGF, Eotaxin, FGF-2, Flt-3, Fractalkine, G-CSF, GM-CSF, GRO-α, IFNα2, IFNγ, IL-1α, IL-1β, IL-1ra, IL-2, IL-3, Il-4, IL-5, IL-5, IL-6, IL-7, IL-8, IL-9, IL-10, IL-12 (p40), IL-12 (p70), IL-13, IL-15, IL-17A, IL-17E/IL-25, IL-17F, IL-18, IL-22, IL-27, IP-10, MCP-1, MCP-3, M-CSF, MDC, MIG, MIP-1α, MIP-1β, PDGF-AA, PDGF-AB/BB, RANTES, TGF-α, TNF-α, TNF-β, and VEGF.

### Flow cytometry

Peripheral blood mononuclear cells were isolated from peripheral blood using Lymphoprep density gradient (STEMCELL Technologies, Vancouver, Canada). Aliquots of cells were frozen in media that contained 90% fetal bovine serum (HyClone, Logan, UT) and 10% dimethyl sulfoxide (Sigma-Aldrich, St. Louis, MO) and stored at <70C. Cells were quick thawed, washed, and incubated with 2% solution of bovine serum albumin (Blocker BSA, ThermoFisher, Waltham, MA) diluted in D-PBS (HyClone) for 5 min. Each sample received a cocktail containing 10 uL Brilliant Stain Buffer (BD Biosciences, Franklin Lakes, NJ), 5 uL True-Stain Monocyte Blocker (BioLegend, San Diego, CA), and the following surface marker antibodies: anti-CD19 (PE-Dazzle594), anti-CD3 (APC), anti-CD16 (Alexa700), HLA-DR (APC/Fire750), and anti-CTLA-4 (PE-Cy7). The following antibodies were then added to each tube individually: anti-CD8 (BUV496), anti-CD4 (BUV661), anti-CD45 (BUV805), anti-CD103 (BV421), anti-TIM3 (BV605), anti-CD56 (BV650), anti-LAG-3 (BV711), anti-CD14 (BB785), and anti-PD-1 (BB700), followed by a 30 min. incubation in the dark at room temperature. Cells were washed once with 2% BSA solution before fixation and permeabilization. Cells were fixed and permeabilized in a one-step reaction with 1X incellMAX (IncellDx, San Carlos, CA) at a concentration of 1 million cells per mL and incubated for 60 min. in the dark at room temperature. Cells were washed once with 2% BSA solution, and analyzed on a Cytoflex LX with 355nm (20mW), 405nm (80mW), 488nm (50mW), 561nm (30mW), 638nm (50mW), 808nm (60mW) lasers (Beckman Coulter Life Sciences, Indianapolis, IN). Analysis was performed with Kaluza version 2.1 software. The panel used in this study is shown in Supplementary Table 1 and examples of the gating strategy is shown in Supplementary Fig. 5.

### CCR5 receptor occupancy

Because CCR5 is a highly regulated receptor especially in infection, inflammation, and cancer, we determined CCR5 receptor occupancy by leronlimab by using phycoerythrin-labeled leronlimab (IncellDx, Inc) in a competitive flow cytometry assay. CCR5-expressing immune cells including CD4+, CD45RO+ T-lymphocytes, CD4+, FoxP3+ T-regulatory cells, and CD14+, CD16+ monocytes/macrophages were included in the panel using the appropriate immunophenotypic markers for each population in addition to PE-labeled leronlimab. Cells were incubated for 30 min. in the dark at room temperature and washed twice with 2% BSA solution before flow acquisition on a 3-laser CytoFLEX fitted with 405nm (80mW), 488nm (50mW), 638nm (50mW) lasers (Beckman Coulter Life Sciences, Indianapolis, IN Life Sciences, Indianapolis, IN). Receptor occupancy was determined by the loss of CCR5 detection over time in these subpopulations (Supplementary Figure 6) and calculated with the following equation:

1-A/B X 100 where A is Day 0 and B is Day 7.

### Measurement of plasma SARS-CoV-2 viral loads

The QIAamp Viral Mini Kit (Qiagen, Catalog #52906) was used to extract nucleic acids from 300–400 μL from plasma sample according to instructions from the manufacturer and eluted in 50 μL of AVE buffer (RNase-free water with 0.04% sodium azide). The purified nucleic acids were used immediately with the Bio-Rad SARS-CoV-2 ddPCR Kit (Bio-Rad, Hercules, CA). Each batch of samples extracted comprised positive and extraction controls which are included in the kit, as well as a no template control (nuclease free water). The Bio-Rad SARS-CoV-2 ddPCR Test is a reverse transcription (RT) droplet digital polymerase chain reaction (ddPCR) test designed to detect RNA from SARS-CoV-2. The oligonucleotide primers and probes for detection of SARS-CoV-2 are the same as those reported by CDC and were selected from regions of the viral nucleocapsid (N) gene. The panel is designed for specific detection of the 2019-nCoV (two primer/probe sets). An additional primer/probe set to detect the human RNase P gene (RP) in control samples and clinical specimens is also included in the panel as an internal control. The Bio-Rad SARS-CoV-2 ddPCR Kit includes these three sets of primers/probes into a single assay multiplex to enable a one-well reaction. RNA isolated and purified from the plasma samples (5.5 μL) were added to the mastermix comprised of 1.1 μL of 2019-nCoV triplex assay, 2.2 μL of reverse transcriptase, 5.5 μL of supermix, 1.1 μL of Dithiothreitol (DTT) and 6.6 μL of nuclease-free water. Twenty-two microliters (22μl) from these sample and mastermix RT-ddPCR mixtures were loaded into the wells of a 96-well PCR plate. The mixtures were then fractionated into up to 20,000 nanoliter-sized droplets in the form of a water-in-oil emulsion in the QX200 Automated Droplet Generator (Bio-Rad, Hercules CA). The 96-well RT-ddPCR ready plate containing droplets was sealed with foil using a plate sealer and thermocycled to achieve reverse transcription of RNA followed by PCR amplification of cDNA in a C1000 Touch thermocycler (Bio-Rad, Hercules CA). Subsequent to PCR, the plate was loaded into the QX200 Droplet Reader (Bio-Rad, Hercules CA) and the fluorescence intensity of each droplet was measured in two channels (FAM and HEX). The Droplet Reader singulates the droplets and flows them past a two-color fluorescence detector. The detector reads the droplets to determine which contain target (positive) and which do not (negative) for each of the targets identified with the Bio-Rad SARS-CoV-2 ddPCR Test: N1, N2 and RP. The fluorescence data is then analyzed by the QuantaSoft 1.7 and QuantaSoft Analysis Pro 1.0 Software to determine the presence of SARS-CoV-2 N1 and N2 in the specimen.

Bio-Rad SARS-CoV-2 RT-ddPCR Thermal Cycling Protocol.

**Bio-Rad SARS-CoV-2 RT-ddPCR Thermal Cycling Protocol**
| **Cycling Step** | **Temperature (°C)** | **Time** | **Number of Cycles** |
| --- | --- | --- | --- |
| Reverse Transcription | 50 | 60 minutes | 1 |
| PCR enzyme activation | 95 | 10 minutes | 1 |
| Template Denaturation | 94 | 30 seconds | 40 |
| Annealing / Extension | 55 | 60 seconds |  |
| Droplet Stabilization | 4 | 30 minutes | 1 |
| Hold (optional) | 4 | Overnight | 1 |

### Statistical Analysis

The inflammatory cytokines IL-1β, IL-6, IL-8, CCL5 levels between groups were compared using non-parametric Kruskal-Wallis test followed by Dunn’s multiple comparison correction to control the experimental wise error rate. To assess reversal of immune dysfunction and CCR5 receptor occupancy as well as cytokine and chemokine levels in severe COVID-19 patients after Leronlimab, Kruskal-Wallis test with Dunn’s multiple comparison correction was used. Changes in SARS-CoV-2 plasma viral loads were assessed using the Mann-Whitney test.

### Patient samples and IRB

All patients were enrolled in this study under an individual patient emergency use investigation new drug (EIND) via FDA emergency use authorization (EUA). The FDA assigned an EIND number for each patient and thus registration in a clinical trial registration agency is not applicable. Informed consent was obtained from patient or their legally authorized representative per 21 CFR Part 50. The Albert Einstein College of Medicine Institution Review Board (IRB) reviewed and approved this study. The IRB was notified within 5 business days of treatment initiation. Within 15 business days of FDA emergency use authorization, Form FDA 3926 along with the treatment plan and the letter of authorization from CytoDyn was submitted to FDA. One 8 mL EDTA tube and one 4 mL plasma preparation (PPT) tube were drawn by venipuncture at Day0 (pre-treatment), Day 3, Day 7, Day 14 post-treatment. Blood was shipped overnight to IncellDX for processing and analysis. Peripheral blood mononuclear cells were isolated from peripheral blood using Lymphoprep density gradient (STEMCELL Technologies, Vancouver, Canada). Aliquots of cells were frozen in media that contained 90% fetal bovine serum (HyClone, Logan, UT) and 10% dimethyl sulfoxide (Sigma-Aldrich, St. Louis, MO) and stored at <70C.

### 10X Genomics 5’ Single-cell RNA-Sequencing

Cryopreserved PBMC cells were thawed in RMPI 1640 complete medium, washed in PBS BSA 0.5%, and cell number and viability measured using a Countess II automated cell counter (Thermo Fisher Scientific). Cells were then diluted to a concentration of 1 million cells per ml for loading into the 10X chip. Single-cell RNA-Sequencing library preparation occurred with the Chromium Next GEM Single Cell Immune Profiling (v.1.1 Chemistry) according to manufacturer’s protocols on a Chromium Controller instrument. The library was sequenced using a High Output Flowcell and Illumina NextSeq 500 instrument. For data processing, Cellranger (v.3.0.2) mkfastq was applied to the Illumina BCL output to produce FASTQ files. Cellranger count was then applied to each FASTQ file to produce a feature barcoding and gene expression matrix. Cellranger aggr was used to combine samples for merged analysis. For quality control, we applied the *Seurat* package for cell clustering and differential expression analyses.

## Data Availability

All primary data presented in this study are available from the corresponding author upon reasonable request. Primary data exists for all figures.

## ACKNOWLEDGMENTS

We gratefully acknowledge the patient participants and their caregivers Scott A Scheinin, Enver Aklin, Magdalena Mamczur-Madry, Sana Ahmed, Pamela Philllippsborn, Vagish Hemmige, Reena Joseph, and Jasmine Thallipllill who made this work possible. We acknowledge Lawrence Drew, Shaheed Abdulhaqq, Justin Greene, Whitney Weber, Jason Reed, Cleiton Pessoa, Katherine Bateman, and Jason Reed for review of the manuscript. This work was supported in part by R01 AI129703 to JBS.

## AUTHOR CONTRIBUTIONS

BKP, HS, KD, KK, JL, SK, NP, JBS conceived of the study, HS and EA coordinated patient care, BKP, HS, AP, EBF, HR, WR, AL, LK, MH, and EH acquired data, BKP, MJC, APSP, CS, BF, HLW, GMW, BSP, SK, JL, AL, LK, MH, EH, LCN, and NP analyzed data, and BKP, MJC, KD, JL, HLW, GMW, BSO, SK, NP, LCN, and JBS wrote the manuscript.

## REFERENCES

1. World Health Organization. Coronavirus disease (COVID-2019) situation reports. https://www.who.int/emergencies/diseases/novel-coronavirus-2019/situation-reports.

2. Huang, C. et al. Clinical features of patients infected with 2019 novel coronavirus in Wuhan, China. Lancet 395, 497–506 (2020).

3. Zhang, D. et al. COVID-19 infection induces readily detectable morphological and inflammation-related phenotypic changes in peripheral blood monocytes, the severity of which correlate with patient outcome. *medRxiv* 2020.03.24.20042655 (2020). doi:10.1101/2020.03.24.20042655

4. Mehta, P. et al. COVID-19: consider cytokine storm syndromes and immunosuppression. Lancet 395, 1033–1034 (2020).

5. Qin, C. et al. Dysregulation of immune response in patients with COVID-19 in Wuhan, China. Clin. Infect. Dis. (2020). doi:10.1093/cid/ciaa248

6. Channappanavar, R. & Perlman, S. Pathogenic human coronavirus infections: causes and consequences of cytokine storm and immunopathology. Semin Immunopathol 39, 529–539 (2017).

7. Nicholls, J. M. et al. Lung pathology of fatal severe acute respiratory syndrome. Lancet 361, 1773–1778 (2003).

8. Law, H. K. W. et al. Chemokine up-regulation in SARS-coronavirus-infected, monocyte-derived human dendritic cells. Blood 106, 2366–2374 (2005).

9. Yen, Y.-T. et al. Modeling the early events of severe acute respiratory syndrome coronavirus infection in vitro. Journal of Virology 80, 2684–2693 (2006).

10. Jacobson, J. M. et al. Antiviral activity of single-dose PRO 140, a CCR5 monoclonal antibody, in HIV-infected adults. J. Infect. Dis. 198, 1345–1352 (2008).

11. Jacobson, J. M. et al. Anti-HIV-1 activity of weekly or biweekly treatment with subcutaneous PRO 140, a CCR5 monoclonal antibody. J. Infect. Dis. 201, 1481–1487 (2010).

12. Jacobson, J. M. et al. Phase 2a study of the CCR5 monoclonal antibody PRO 140 administered intravenously to HIV-infected adults. Antimicrob. Agents Chemother. 54, 4137–4142 (2010).

13. Dhody, K. et al. PRO 140, a monoclonal antibody targeting CCR5, as a long-acting, single-agent maintenance therapy for HIV-1 infection. HIV Clin Trials 19, 85–93 (2018).

14. Olson, W. C. et al. Differential inhibition of human immunodeficiency virus type 1 fusion, gp120 binding, and CC-chemokine activity by monoclonal antibodies to CCR5. Journal of Virology 73, 4145–4155 (1999).

15. Akalin, E. et al. Covid-19 and Kidney Transplantation. N Engl J Med NEJMc2011117 (2020). doi:10.1056/NEJMc2011117.

16. Richardson, S. et al. Presenting Characteristics, Comorbidities, and Outcomes Among 5700 Patients Hospitalized With COVID-19 in the New York City Area. JAMA (2020). doi:10.1001/jama.2020.6775

17. Yu, T-M et al. RANTES mediates kidney ischemia reperfusion injury through a possible role of HIF-1α and LncRNA PRINS. Scientific Reports (2016) 10.1038/srep18424

18. Chen, L. Functional roles of CCL5/RANTES in liver disease. Liver Research (2020) 28e34

19. Lescure, F.-X. et al. Clinical and virological data of the first cases of COVID-19 in Europe: a case series. Lancet Infect Dis (2020). doi:10.1016/S1473-3099(20)30200-0

20. Halama, N., et al. Tumoral immune cell exploitation in colorectal cancer metastases can be targeted effectively by anti-CCR5 therapy in cancer patients. Cancer Cell (2016) 29, 587–601, 2016.

21. Grillet, F., Behr, J., Calame, P., Aubry, S. & Delabrousse, E. Acute Pulmonary Embolism Associated with COVID-19 Pneumonia Detected by Pulmonary CT Angiography. Radiology 201544 (2020). doi:10.1148/radiol.2020201544

22. Machlus, K. R. et al. CCL5 derived from platelets increases megakaryocyte proplatelet formation. Blood 127, 921–926 (2016).

23. Coronavirus survivor credits artificial antibody experimental treatment for recovery. Los Angeles CBS Local (2020). https://losangeles.cbslocal.com/2020/04/10/coronavirus-survivor-leronlimab/

24. US National Library of Medicine. Study to Evaluate the Efficacy and Safety of Leronlimab for Mild to Moderate COVID-19. ClinicalTrials.gov. https://clinicaltrials.gov/ct2/show/NCT04343651 (2020).

25. US National Library of Medicine. Study to Evaluate the Efficacy and Safety of Leronlimab for Patients With Severe or Critical Coronavirus Disease 2019 (COVID-19). ClinicalTrials.gov. https://clinicaltrials.gov/ct2/show/NCT04347239 (2020).

